# Hidden Burden of a Measles Outbreak Revealed by Genomic and Transmission Models

**DOI:** 10.64898/2026.07.09.26357695

**Authors:** Abbey Marye, Damon J.A. Toth, George Vega Yon, Jake Wagoner, Eric T Lofgren, Matthew H. Samore, Andrew T. Pavia, Leisha D. Nolen, Kenneth Komatsu, Kelly Oakeson, Lindsay T. Keegan

## Abstract

Declining childhood vaccination rates have fueled a resurgence of measles in the United States. Surveillance systems may not accurately measure the true extent of outbreaks. As of May 2026, the largest ongoing measles outbreak in the United States originated along the Utah-Arizona border in a community with high vaccine exemption rates and limited engagement with healthcare systems, leading to incomplete testing and reporting. To quantify the true outbreak size, we used two independent approaches with complementary data sources: a phylodynamic analysis and an agent-based model. Both methods found significant underreporting, estimating the true outbreak size to be 3.1- to 4.8-fold larger than reported, with confirmed cases representing only 20.96%-32.5% total infections. These findings suggest that substantial underreporting of measles occurs, especially in tight knit communities. The use of complementary analytical approaches to evaluate completeness of reporting can reveal the extent of measles transmission and aid control efforts.

## Main

Once a common childhood infectious disease infecting 3-4 million people in the United States (US) annually^1^, measles was declared eliminated in 2000 following the introduction and scale up of an effective vaccine^2^. However, because measles is highly transmissible (R_0_:12-18)^3^, very high vaccination coverage is required to prevent transmission (≥97%), and even small pockets of inadequate vaccination can sustain outbreaks^2,4,5^. On average, rates of measles, mumps, and rubella (MMR) vaccination coverage have historically remained high across the US, however, vaccine hesitancy has increased following the COVID-19 pandemic, resulting in pockets of lower coverage, susceptible to outbreaks^6,7^. Since 2014, measles outbreaks in the US have become increasingly large and sustained, with 2,288 cases reported in 2025^5,8,9^.

As of May 2026, the largest active measles outbreak in the US continues in Utah, originating from an outbreak along the Utah-Arizona border^10^. The outbreak began in a tight-knit community, straddling the border, in a region with historically low childhood vaccination rates and limited engagement with public health systems, largely due to vaccine skepticism and government mistrust^11^. Early in the outbreak, clinicians and public health officials were told of individuals with symptoms consistent with measles who did not seek care, and therefore were never tested or reported, suggesting that surveillance data may not fully capture the extent of transmission^10^. State-level MMR vaccination coverage in Utah is below the national average (Figure 1a), and vaccine exemption rates are the second highest in the country (10.3% in 2025). Arizona’s vaccine exemption rate of 8.5% is similarly well above the national average of 3.3%^12,13^. The Utah-Arizona border outbreak spans Washington County, Utah, and Mohave County, Arizona, where kindergarten MMR coverage declined from 92.9% to 78.4% and from 93.3% to 86.7%, respectively, between the 2015-2016 and 2024-2025 school years (Figure 1b)^14^. The affected community has even lower vaccination coverage than the surrounding counties (Figure 1a) and is characterized by larger-than-average household sizes and a younger than average population, factors which may facilitate transmission once infection is introduced^15^.

**Figure 1:**
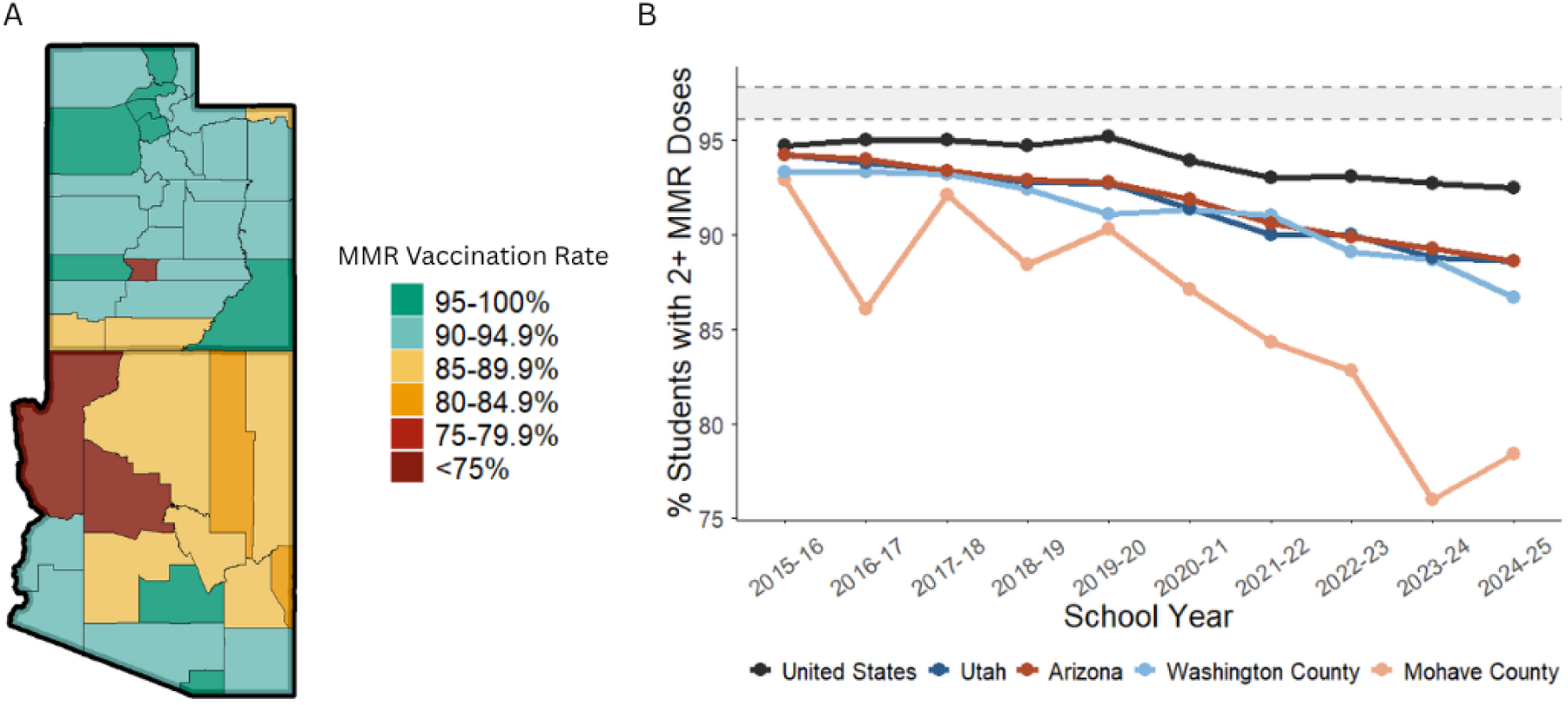
Spatial and temporal patterns of MMR vaccination coverage in the Utah-Arizona Border Outbreak region. A) Map of MMR vaccination coverage by county for all students in Utah and for kindergarteners in Arizona. **B)** Percent of _≥_2 MMR vaccination doses over ten school years (2015 -2025) for kindergarteners in the US, Arizona, and Mohave County, Arizona and for all students in Utah and Washington County, Utah; the critical vaccination threshold (96.1%-97.8%) is indicated by the shaded bar.

Measles surveillance has historically depended on passive case detection through clinical diagnosis. Reporting completeness in the US has historically been low, estimated to range from 22-67%^16,17^. More recently, many jurisdictions have shifted towards requiring laboratory-confirmation, which may improve diagnostic specificity in exchange for significant undercounting, but completeness of reporting for laboratory-confirmed measles has not been explored. Socioeconomic and structural factors that drive pockets of low vaccination coverage may also impact healthcare-seeking behavior and access to diagnostic testing, contributing to under-reporting in these communities^6,7^. Recent wastewater surveillance studies have detected measles virus before reported cases and in counties without reported cases, suggesting that cases are occurring in these areas that are undetected^15^. Accurate estimation of outbreak size is essential for public health response. Concerned that not all cases were being detected by routine surveillance, we sought to estimate the true outbreak size and quantify the degree of under-ascertainment using two independent approaches and complementary data sources^18^.

## Results

### Outbreak Description

The outbreak originated in August 2025 in Mohave County, Arizona, with rash onset identified in an unvaccinated individual for whom no source of exposure could be established. Subsequent cases meeting the epidemiologic criteria for outbreak classification were reported over the ensuing days. The first epidemiologically linked case in Washington County, Utah was identified approximately two weeks after initial outbreak detection. Sporadic measles cases reported in Utah and Arizona prior to this outbreak did not have any epidemiologic link to the current cluster. Among cases linked to the Utah-Arizona border outbreak, 96.8% occurred in unvaccinated individuals or individuals with unknown vaccination status, compared with 93% of cases reported nationally during the same period (Figure 2a). Although the affected community has a younger age distribution relative to the US population (Table SI 1), the age distribution of cases in this outbreak shifted toward older age groups compared to cases reported elsewhere in the United States (Figure 2b). The primary outbreak along the Utah-Arizona border began to subside by February 28,2026 and measles has since spread throughout Utah (Figure 2c, d).

**Figure 2:**
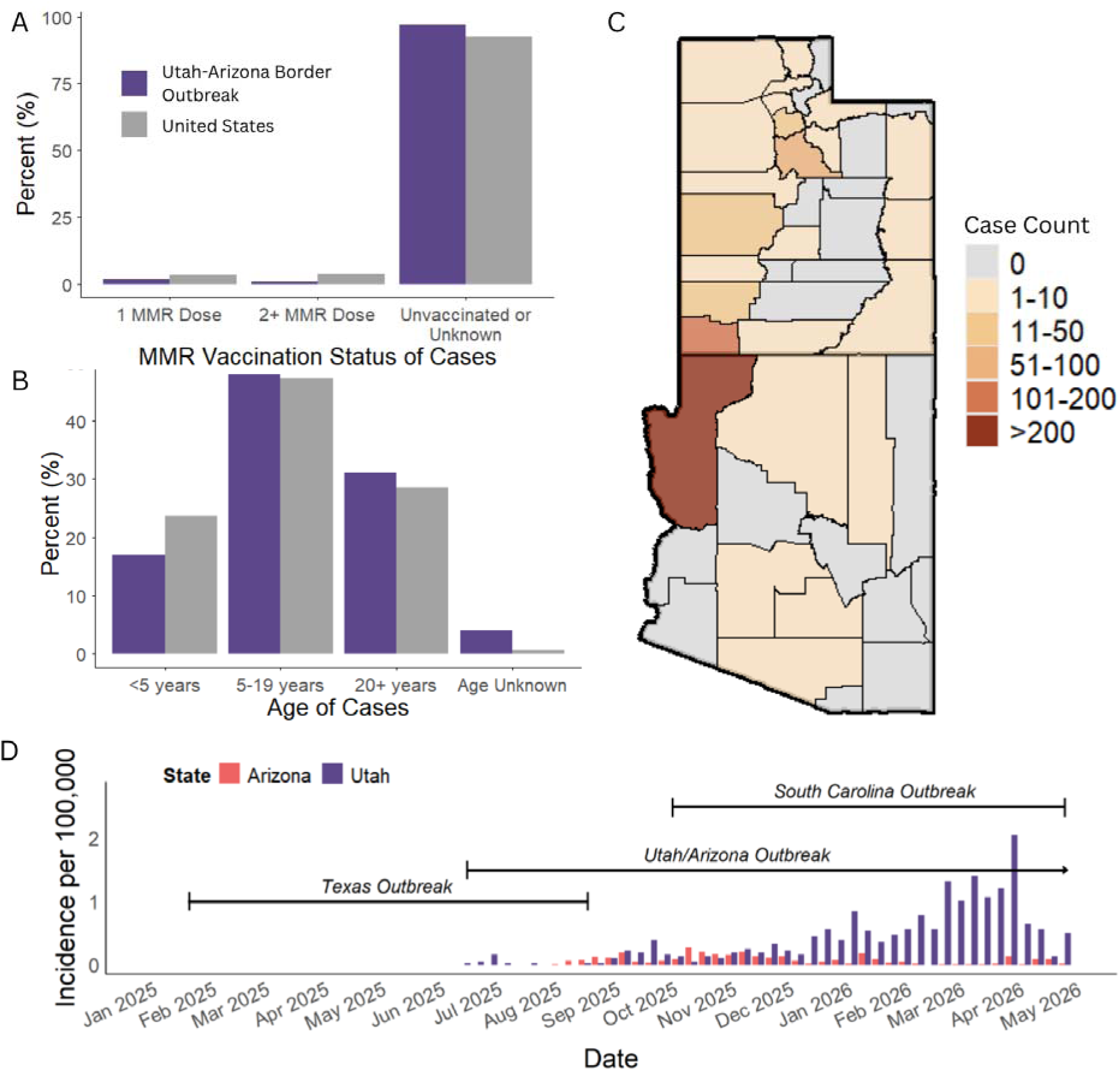
Characteristics and Geographic Spread of the 2025-2026 Utah-Arizona Border Outbreak in national context. A) MMR vaccination status and B) age distribution of cases in the Utah-Arizona border outbreak compared to all other reported U.S. cases. C) Map of confirmed cases in Utah and Arizona, July 2025 to May 2026. D) Timing and magnitude of the Utah-Arizona border outbreak, relative to other major outbreaks contributing to the 2025-2026 US measles outbreaks.

### Phylodynamic Estimation of Effective Population Size and Outbreak Size

To estimate effective population size over time, we performed a coalescent-based phylodynamic analysis for all infections linked to the Utah-Arizona border outbreak, using whole-genome sequence (WGS) data from clinical isolates. This approach infers viral population size from the timing and genetic relationships among sampled sequences, capturing all infections connected to the outbreak through shared transmission chains regardless of geographic location. Because it cannot resolve where inferred but unsampled infections occurred, this estimate of the outbreak size is not restricted to the affected community but encompasses all linked transmission across Utah, Arizona, Idaho, and Colorado. Connection to the outbreak was defined using two criteria: cases were considered epidemiologically linked if they had verified contact with a confirmed outbreak case, and genomically linked if their sequences were phylogenetically related to a confirmed Utah-Arizona border outbreak case; neither criterion restricted inclusion by location. Analyses were restricted to cases occurring before February 28, 2026, to focus on the primary outbreak wave within the affected community.

Measles-positive specimens were collected through routine surveillance during the outbreak and sequenced using the ARTIC Measles 400-bp v1.0.0 primer scheme on the Illumina NextSeq platform^19,20^. Sequences meeting pre-specified quality thresholds were included in downstream analyses. These sequences were used to estimate the effective population size over time using a coalescent-based extended Bayesian skyline model in BEAST2 (v2.7.7)^19^ that incorporated sampling times and genetic relationships among sequences, based on the 53% of confirmed cases that were sequenced^21^ (Figure 3). To estimate cumulative infections from the extended Bayesian skyline model, we derived the time-varying infection prevalence from the inferred effective population size by accounting for the reproductive number (R_0_ = 9) heterogeneity in transmission (k = 0.34)^22–24^. We then integrated these prevalence estimates over the course of the outbreak to obtain cumulative incidence and calculate 95% credible intervals (95% CrI)^25,26^.

**Figure 3:**
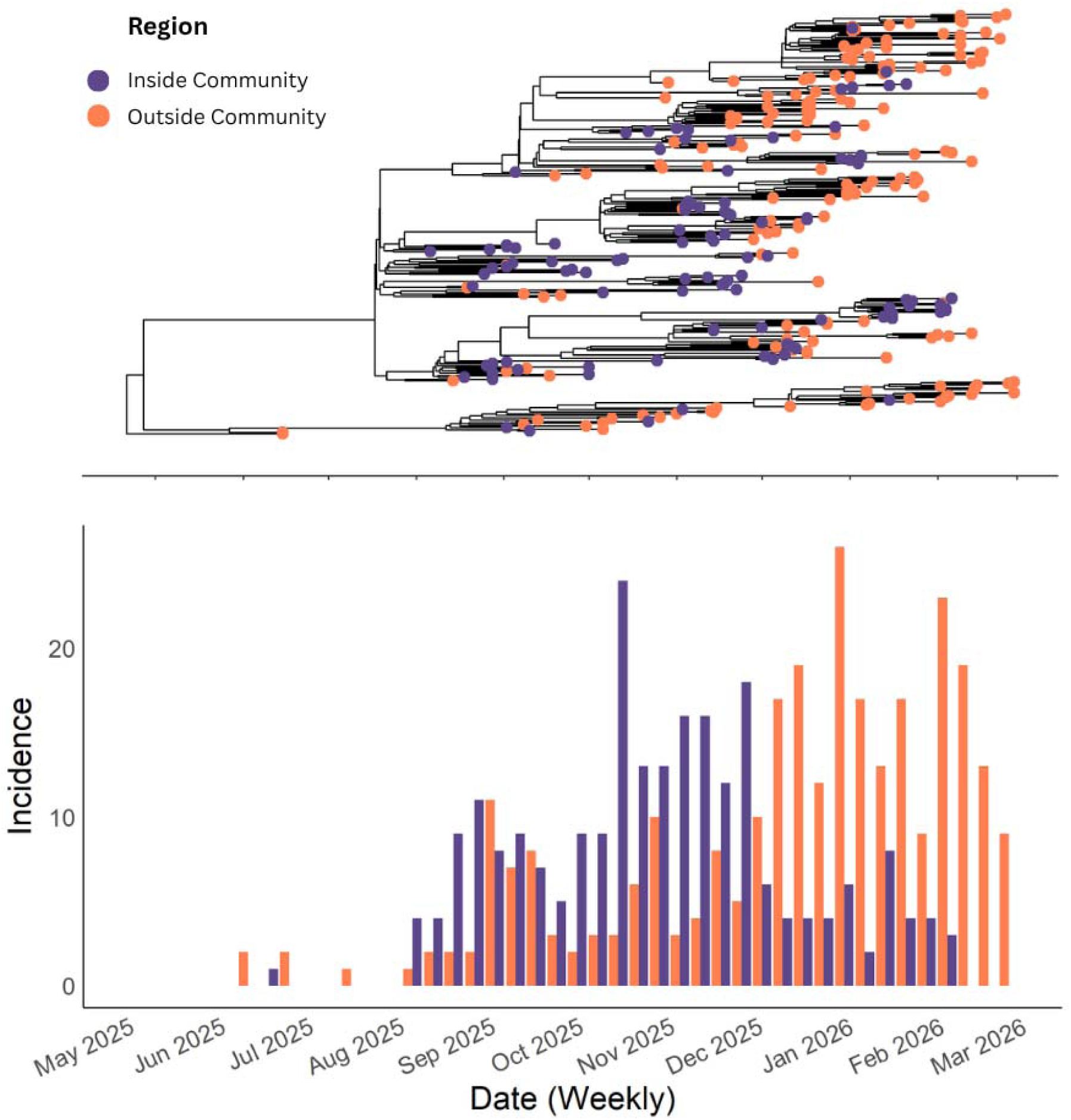
Genomic relationships and reported incidence among confirmed cases linked to the Utah-Arizona Border Outbreak. Phylogenetic tree of the cases linked to the Utah-Arizona border outbreak. Purple nodes are cases in the affected community whereas orange nodes are linked cases outside of the affected community. Below is a bar plot of the reported incidence in the affected community (purple) and the cases linked to the Utah-Arizona border outbreak, but not part of the affected community (orange).

We estimated the total number of cases epidemiologically or genomically linked to the Utah-Arizona border outbreak to be 1,798 infections (95% CrI, 1,050 to 3,573 infections, and 2,051 infections (95% CrI, 1,300 to 3,885 infections), respectively (Figure 4a,b). No single reported case count serves as an ideal denominator for estimating under-ascertainment because the phylodynamic analysis captures a transmission network that cannot be precisely bounded geographically. As such, we compare our estimates to available case counts to provide a plausible range for underreporting. Comparing estimates to their most analogous reported case counts, we find the epidemiologically linked estimate was 4.12-fold higher than epidemiologically linked reported cases (n=436), and the genomically linked estimate was 3.93-fold higher than genomically linked reported cases (n=522). When compared to total confirmed cases in Utah and Arizona (n=585), the epidemiologically and genomically linked estimates represented 3.1-fold and 3.5-fold under-ascertainment, respectively. These estimates suggest that across the 95% credible intervals, reported cases represented approximately 15.1% to 55.7% of total infections.

**Figure 4:**
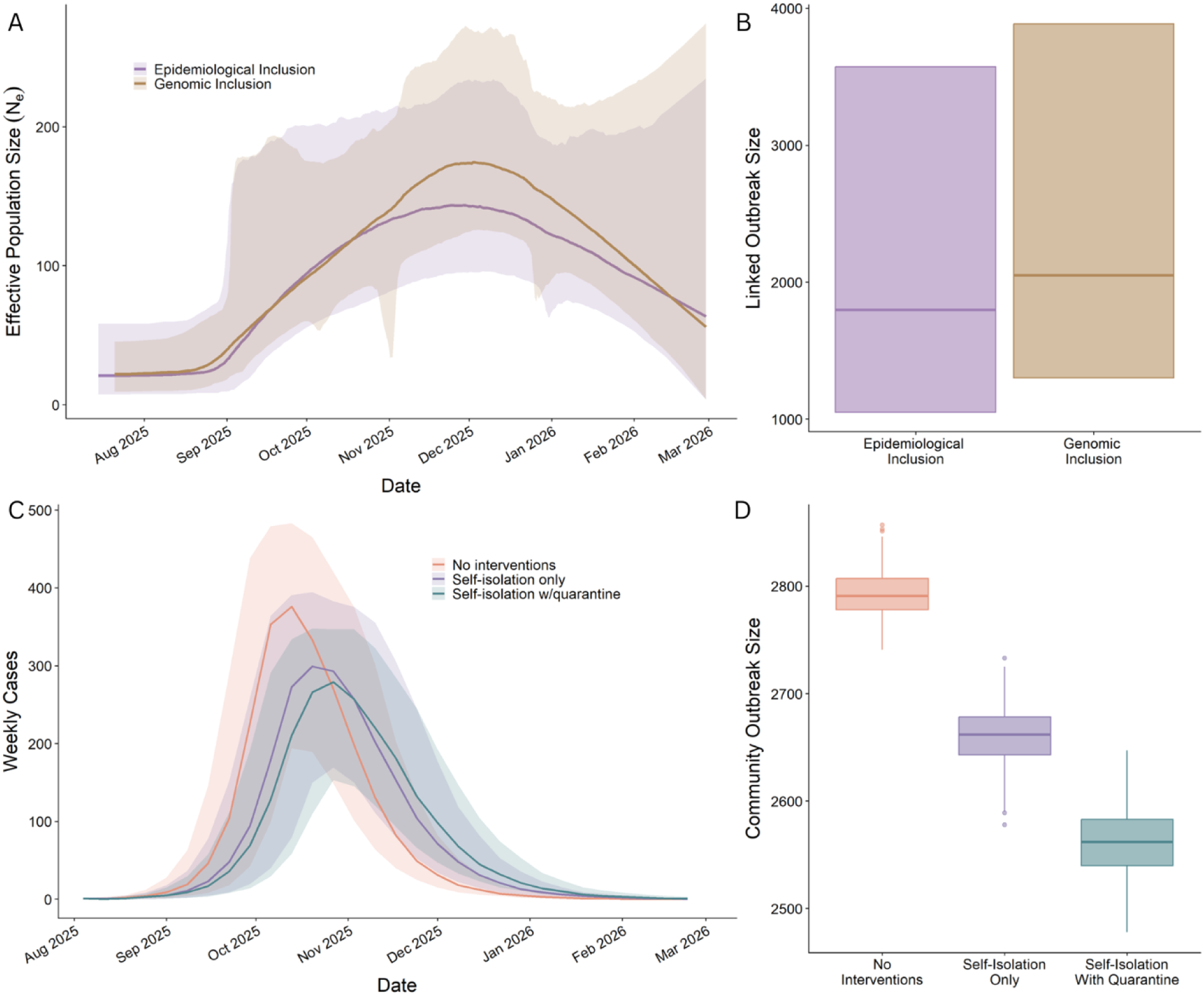
Estimates of Effective Population Size during the Utah-Arizona Border Measles Outbreak. A) Effective population size estimates from phylodynamic estimation are plotted over time starting from June 15, 2025, to February 28, 2026 (latest sampling date) for estimates from the epidemiological inclusion and the genomic inclusion. Central posterior densities are shaded in purple and gold for epidemiological and genomic inclusion, respectively. B) A boxplot showing the total outbreak size estimation from phylodynamic estimation for both epidemiological inclusion (purple) and genomic inclusion (gold). C) Estimated number of infections from the agent-based model plotted over time. Median (line) and interquartile range (shaded region) for estimates accounting for no interventions (red), self-isolation without quarantine (purple), and self-isolation with quarantine (green). D) A boxplot showing the total outbreak size estimation from the agent-based model for no interventions (red), self-isolation without quarantine (purple), and self-isolation (green).

The phylodynamic analysis estimated that the outbreak peaked in December 2025 (Figure 4a) whereas reported cases in the affected community peaked in November 2025 (Figure 3b). The trajectory estimated by the phylodynamic analysis reflects the aggregate signal of two partially overlapping, phase-shifted transmission waves: one wave within the community of interest, and another in other areas of Utah (Figure SI 6).

### Agent-Based Simulation of Outbreak Size

To estimate outbreak size specifically within the affected community we developed a stochastic, metapopulation agent-based model incorporating age-structured contact patterns and school-based mixing, implemented using the epiworldR^27,28^ and measles R packages^29^. The model uses a modified SEIR structure that includes a prodromal infectious stage prior to rash onset, a rash stage, and hospitalization. In addition to disease dynamics, we modeled three mechanisms of transmission reduction: spontaneous contact reduction following rash onset (“self-isolation”), isolation of confirmed cases, and quarantine of identified contacts, each parameterized by an independent adherence probability (Figure SI3). Model parameters were derived from empirical data where available; key assumptions were evaluated through sensitivity analyses

Population size and age structure were derived from the 2024 U.S. Census (n = 5,913)^30^. Vaccination coverage was estimated from school-level data obtained from the Utah Department of Health and Human Services and the Arizona Department of Health Services, with coverage for non-school-age adults estimated from published national estimates; overall vaccination coverage in the affected community was estimated at 50.1%. Contact patterns were specified using age-structured mixing matrices from the POLYMOD study^31^; robustness to this assumption was evaluated in a sensitivity analysis.

To calibrate the transmission rate, we estimated the time-varying effective reproduction number (Rₜ) from publicly available Utah case data using a maximum-likelihood approach during the initial exponential growth period in August 2025^2,32^. From Rₜ, we derived R_0_ (R_0_ = 9; Figure SI 3) and calibrated the model accordingly. We conducted sensitivity analyses across R_0_ = 5-17 to evaluate the robustness of our findings to this assumption.

Although anecdotally, isolation was variable and quarantine not extensively enforced, the dynamics of the reported cases do not reflect unmitigated transmission. It is likely that despite minimal public health directives, individuals infected with measles changed their behavior as a result of their infection. Our model explores different assumptions of isolation and quarantine to reflect this uncertainty. We present three plausible behavioral scenarios in the main text. In the first scenario, we assume individuals do not reduce contacts following illness (“no interventions”). The second scenario we assume individuals reduce mixing by 50% following rash onset without any public health isolation or contact tracing (“self-isolation only”). In the final scenario, we assume individuals reduce mixing by 75% following rash onset and 25% of cases engage with public health contact tracing, resulting in effective quarantine of 90% of identified contacts (“self-isolation with quarantine”), with the latter two representing plausible bounds on behavioral response. Twenty-five possible combinations of self-isolation and public health quarantine adherence are reported in the supplement.

Simulations were initialized with a single infectious individual on August 6, 2025, and 1000 stochastic realizations were performed per scenario. 220 (22.0%), 301 (30.1%), 390 (39.0%) resulted in early extinction (<10 cases) and were excluded from analysis, as the observed outbreak did not fade out, for no interventions, self-isolation only, and self-isolation with quarantine, respectively. Among the remaining simulations, the median outbreak size was 2,791 infections (interquartile range [IQR]: 2,778-2,807 infections) for outbreaks with no interventions, 2,662 infections (IQR: 2,643-2,678 infections) for outbreaks with self-isolation only, and 2,226 infections (IQR: 2,197-2,258 infections) for outbreaks with self-isolation with quarantine (Figure 4d). These estimates are 4.8-fold, 4.6-fold, and 3.8-fold higher respectively than the confirmed case count of 585 confirmed case count in Utah and Arizona as of February 28, 2026. Thus, reported cases represent approximately 20.96% 21.98%, and 26.28% of infections depending on the level of control from behavior change. Simulations with interventions yielded substantially smaller outbreaks. The probability of a large outbreak (>100 cases) was 78.0%, 69.9% and 60.6% for outbreaks with no interventions, isolation without quarantine, and isolation with quarantine respectively.

The median peak incidence week occurred in October and early November 2025, for simulations with no interventions, October 13, 2025, with 562 infections, for simulations with self-isolation only, and October 27, 2025, with 482 infections, for simulations with self-isolation and quarantine, November 6, 2025, with 311 infections (Figure 4c). The phylodynamic estimates of outbreak size were not substantially larger than those from the agent-based simulations, reflecting concordance between approaches.

In sensitivity analyses across a range of basic reproduction numbers (R_0_ = 5-17), median outbreak size among simulations that did not result in early extinction, the impact was modest. Estimates ranged from 2, 230 infections (IQR: 2,202-2,256 infections at R_0_ = 5 to 2,989 infections (IQR: 2,980-2,997 infections) at R_0_ = 17 for simulations without interventions, 1,915 infections (IQR: 1,882-1,948 infections at R_0_ = 5 to 2,955 infections (IQR: 2,946-2,964 infections) at R_0_ = 17 for simulations with self-isolation without quarantine, and 1,344 infections (IQR: 1,274 -1,401 infections) at R_0_ = 5 to 2,668 infections (IQR: 2,649-2,684 infections) at R_0_ = 17 for simulations with self-isolation and quarantine (Table SI3). Peak timing also varied with R_0_, ranging from September 22, 2025, to January 26, 2026, depending on assumptions of R_0_ and interventions (Table SI 4).

## Discussion

Estimating the true size of a measles outbreak is a long-standing challenge that limits the ability to understand the true impact^16,17^. As of May 2026, the largest active measles outbreak in the US was ongoing on the Utah-Arizona border with more than 600 reported cases. The outbreak originated in a community with low vaccination coverage and limited engagement with healthcare systems and spread throughout Utah. Our findings demonstrate that in the modern era where the availability of PCR testing and electronic laboratory reporting might be expected to provide more complete reporting than passive surveillance of clinical cases, the true size of a measles outbreak is not captured by surveillance. Using two independent approaches based on complementary data sources, we estimate that the Utah-Arizona border outbreak was substantially larger than reported, with confirmed cases representing approximately 20.96%-32.5% of total infections. Importantly, while both approaches identify substantial under-ascertainment, they do not estimate the same underlying quantity. The phylodynamic analysis estimated between 1,798 and 2,051 infections linked to the Utah-Arizona border outbreak (both within and outside the originating community), depending on case inclusion criteria. Similarly, the agent-based model estimated a median of 2,226 to 2,791 infections within the affected community, depending on assumptions of interventions. These differences reflect the populations captured by each approach: the phylodynamic analysis includes all infections linked to the outbreak regardless of location, whereas the agent-based model is bounded by the affected community. Phylogenetic reconstruction indicates that transmission moved frequently between the affected community and surrounding areas in both directions (Figure 3), consistent with the broader scope of the phylodynamic estimate. Despite these differences, the credible interval from the phylodynamic analysis encompassed the estimates from the agent-based model, providing strong evidence for substantial under-ascertainment of infections during the outbreak. The convergence of estimates from two methods relying on independent data sources and distinct modeling assumptions strengthens confidence that the degree of under-ascertainment is not just an artifact of model assumptions.

For a case to be included in surveillance, a patient must have symptoms, seek healthcare, have the correct diagnosis made, and that diagnosis be reported to public health. In routine surveillance this limitation is well recognized but often difficult to quantify. During a large-scale outbreak, however, the degree of under-ascertainment has direct consequences for public health response: estimates of transmission intensity, assessments of intervention effectiveness, and projections of outbreak trajectory all depend on case counts that, if substantially incomplete, will mislead in proportion to their incompleteness. Under-ascertainment also complicates estimation of true disease burden, including rates of severe outcomes, hospitalization, and population-level immunity accrued through infection, each of which informs both the immediate response and longer-term surveillance priorities. A number of methods have been used in the past to examine the completeness of measles surveillance, including household surveys, capture-recapture analysis of hospitalized cases, and examination of administrative data^17^. All have found substantial under-estimation of the true burden. These studies however were conducted when the diagnosis of measles largely relied on a clinical case definition, and reporting was manual. In the current era, most confirmed cases are based on laboratory testing with molecular testing, or less commonly, serology. In many jurisdictions, including Arizona and Utah, positive tests for reportable diseases are automatically transmitted to public health via electronic laboratory reporting. These factors improve the completeness and accuracy of surveillance, however they still depend on individuals presenting for care and being tested. However, using two complementary methods, we found substantial under-ascertainment of measles in this outbreak. The magnitude of under-ascertainment likely varies by setting. Factors such as access to care, health care seeking behavior, testing practices, and the efficiency of laboratory reporting are probable drivers. With many large outbreaks of measles arising in tight knit communities, it is critical to better understand how these factors influence reporting in different settings.

The two approaches we used were not formally integrated and were instead applied independently to different data sources and indeed generated estimates for different underlying populations: the total number of cases related to the outbreak for the phylodynamic models and the total number of cases in the outbreak community for the agent-based model. Recent work has shown that integrating genomic and epidemiologic data can improve inference of outbreak dynamics; however, such approaches typically rely on a single integrated model^33–36^. Such joint frameworks combine pathogen sequence data with epidemiologic observations to reconstruct transmission, estimate reproduction numbers, and infer unobserved infections. However, because these approaches rely on a single integrated model they cannot provide independent validation of findings. Our dual, independent approach draws on distinct data sources and assumptions and therefore allows for independent assessment of outbreak size. Estimates from multiple approaches can lend confidence to inference, particularly in settings where surveillance is believed to be incomplete.

This study has several limitations. The phylodynamic analysis relies on coalescent assumptions, and the sampling proportion is unknown; in a setting with substantial under-ascertainment, observed sequences may not be representative of all infections. Limited genetic diversity in measles viruses over short time scales also constrains phylogenetic resolution. The estimates presented in this study should not be used to determine rates of hospitalization or severe sequelae, since we know that this community provided home treatments to many individuals who would have been hospitalized in other communities.

Similarly, the agent-based model depends on assumptions regarding contact structure, transmission dynamics, and vaccination coverage. Contact patterns were based on age structure and school-based mixing and may not fully capture behavioral heterogeneity, particularly in this unique population. Transmission was calibrated to a fixed basic reproduction number; however, sensitivity analyses across a range of plausible R_0_ values (5-17) yielded similar estimates of outbreak size, suggesting that findings are robust to uncertainty in this parameter. Isolation and quarantine were applied uniformly throughout the simulation upon case detection, and the model did not incorporate post-exposure prophylaxis. Case detection was represented as a fixed probability, which may not capture variability in healthcare-seeking behavior or testing practices across individuals and over the course of the outbreak. The model also assumes no change in behavior over the course of the outbreak, an assumption that may not hold given the substantial public attention this outbreak received. Finally, in the absence of a gold standard for true outbreak size, direct validation of either approach is impossible.

The analytical approaches applied here are pathogen agnostic and are useful in settings where surveillance completeness is uncertain, such as influenza and mpox. However, the magnitude of under-ascertainment in this outbreak reflects characteristics specific to this community, including low vaccination coverage, large household sizes, and limited engagement with healthcare systems, and may not be representative of other settings. In settings with higher vaccination coverage or more diffuse contact patterns, both the magnitude and timing of transmission may differ.

By combining genomic and mechanistic modeling approaches, we conclude that the Utah-Arizona border outbreak was considerably larger than reported. These findings suggest that under-ascertainment may be substantial even in the modern setting with laboratory testing, electronic laboratory reporting, and effective public health infrastructure. Although this outbreak occurred in a close-knit community with limited engagement with external institutions, the mechanisms driving underreporting are unlikely to be unique to it. Vaccine skepticism, distrust of public health authorities, and limited engagement with healthcare systems are increasingly common features of populations with low vaccination coverage in the United States, and the same factors that lead individuals to decline vaccination may also reduce their likelihood of seeking care, accepting testing, or otherwise being captured by passive surveillance when illness occurs. The result is a structural bias in which outbreaks become less visible precisely in the communities where they are most likely to occur.

As pockets of low coverage continue to grow, the need for accurate assessment of the impact will increase. Complementary approaches such as sero-surveillance, wastewater monitoring, or community-based case finding may be needed to accurately characterize burden and inform outbreak response in these settings.

## Online Methods

This study was conducted as part of public health surveillance and outbreak response and was determined not to require IRB review.

### Outbreak Description

#### R_0_ Estimation

We estimated the time-varying effective reproduction number (Rₜ) using the maximum-likelihood based approach of White and Pagano on publicly available Utah case data^32,37^. From Vink et al. 2014^38^, we assumed that the serial interval was given by a gamma distribution with mean 9.9 days and standard deviation 2.5 days. We also explored a mean of 11.7 and 13.8 days. From Rₜ, we estimated R_0_ using:

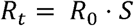

where S is the susceptible fraction. We calculated S as the product of the vaccination coverage in the population and the vaccine effectiveness (assuming 97%)^39^. Since there is considerable uncertainty around the vaccination rate in the population outside of school-aged children, we estimated R_0_ using multiple estimates for vaccination coverage (Figure SI 4):

1. The assumption of adult vaccination used in the agent-based model^40^: where assume that the vaccination rate of 18-24-year-olds was equal to the average of the high schools, the vaccination rate of 25-44-year-olds was 66%, the vaccination rate of 45-69-year-olds was 95% and we assume that all 70+ year olds, had measles as children, giving us a total vaccination coverage of 50.1%.
2. Estimation of adult vaccination based on Washington county^14^: all adults under 70 are equal to the average of the county, which is 78% and we assume that all 70+ year olds, had measles as children, which gives a vaccination coverage of 57.5%
3. Estimation based on Mohave County, Arizona; Utah; and Arizona^8^: all adults under 70 are equal to the average of the county and the two states (86%) and we assume that all 70+ year olds, had measles as children, which gives a vaccination coverage of 61.9%
4. Estimation based on vaccination coverage 10 years ago^14^: all adults under 70 are equal to the average of the two counties from 10 years ago (93%) and we assume that all 70+ year olds, had measles as children. This gives a vaccination coverage of 65.4%

### Phylodynamic Estimation of Effective Population Size and Outbreak Size

#### Whole Genome Sequence Data and Processing

Measles virus PCR-positive clinical specimens submitted to or tested at the Utah Public Health Laboratories (UPHL) were whole genome sequenced using the Artic Measles 400 bp v1.0.0 primer set from Primal Scheme on the Illumina NextSeq platform^20,41^. Raw reads were processed and consensus sequences were generated using the Cecret pipeline^42^. The Cecret pipeline includes quality filtering (seqyclean), alignment (bwa), primer trimming (ivar), and consensus genome generation (ivar). Taxonomic classification was performed using Kraken2 on the reads. Raw sequence data have been deposited in the NCBI Sequence Read Archive under the BioProject assession number PRJNA1293457.

#### Case and Sequence Inclusion

For this analysis, we approached case and sequence inclusion in two different categories, strictly epidemiologic inclusion and genomic inclusion (Figure SI 1).

Cases met the criteria for epidemiologic inclusion if they had a disease onset date between August 6th, 2025, and February 28th, 2026. After February 28, 2026, only four additional cases were reported among members of the affected community, whereas 57 cases were reported in surrounding areas and 210 cases were reported overall across jurisdictions. These patterns indicate that later transmission was primarily occurring outside the initial outbreak community. Cases from Utah, Arizona, Idaho, and Colorado with verifiable epidemiological links were included. However, cases that were genomically distinct from the outbreak were removed since they were reliably ruled out via genomic evidence.

For genomic inclusion, all cases with a disease onset date before February 28th, 2026, were considered. The rationale for this restriction was that subsequent transmission was predominantly observed outside the community, with only 4 cases reported in the Utah-Arizona border outbreak and 210 cases reported across Utah. For genomic inclusion, all cases with onset dates before February 28, 2026, from Utah, Arizona, Idaho, and Colorado were included regardless of whether they pre-dated the outbreak’s recognized start. Cases from genomically distinct clusters, cases with no epidemiologic or genomic link to the Utah-Arizona border outbreak, and sequences that were genomically distinct from all other outbreak sequences were excluded. Furthermore, sequences were required to have ≥90% consensus coverage and ≤25% unknown or missing nucleotides to be included for analysis.

**Figure SI1:**
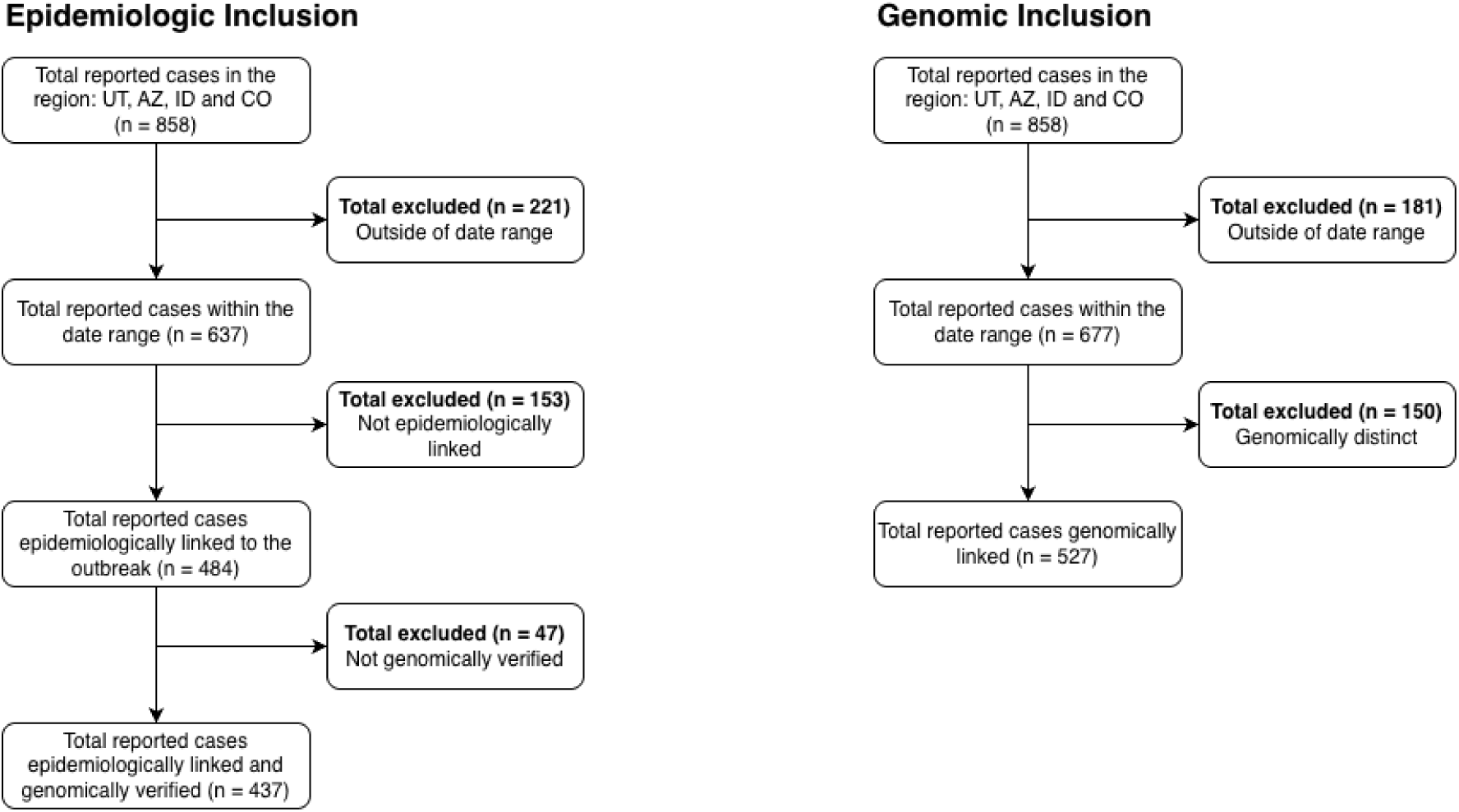
Diagrams of the inclusion criteria for the phylodynamic analysis for. A) epidemiologic inclusion criteria and B) genomic inclusion criteria.

#### Estimation of population size (N_e_)

We estimated effective population size over time ( ) using an extended Bayesian Skyline analysis of the whole genome, rather than just the N450 region of the genome, as it provides greater phylogenetic resolution than the N450 region alone by maximizing the number of informative sites available for reconstruction^43^. Coalescent-based methods are robust to low sampling proportion, making this approach well-suited to settings where a substantial fraction of infections are undetected^44^. By utilizing the timing of samples and the timing of coalescent events between samples, an effective population size can be estimated for each point in time^43^. Each case was assigned a date corresponding to the disease onset date recorded in Utah’s disease surveillance database or the specimen collection date, if the onset date was missing. ClustalW alignments for both inclusion categories were created using clustalo. These alignments were imported into Beauti^45^ along with the case dates. We specified an HKY substitution model^46^, a strict molecular clock with a rate of 0.000661^47^, and used a Coalescent Extended Bayesian Skyline prior to allow effective population size to vary through time. MCMC was run in BEAST2^45^ and R to complete the extended Bayesian Skyline analysis, and posterior distributions were processed using a modified version of plotEBSP.R^21^ to estimate and plot effective population size at each time point.

#### Outbreak size estimation from effective population size (N_e_)

We estimated cumulative outbreak size using two approaches, which we treated as a sensitivity analysis. In the primary approach, we followed the approach of^25,26^ converting (*N_e_*τ) estimates to point prevalence at each time point by accounting for heterogeneity in transmission using I = *N_e_*σ^2^, where σ^2^ is the variance in the offspring distribution, and is given by:

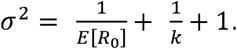

We based on the estimates of R we assumed E[R ] = 9 and used a dispersion parameter of k = 0.34^22–24^. Cumulative incidence was estimated by summing incident infections across sequential generation time windows over the outbreak period.

As a sensitivity analysis, we estimated the outbreak size from the effective population size by dividing by the estimated generation time for measles (11.9 days), plotted N_e_ at each time point using a modified version of plotEBSP.R, and estimated total outbreak size by taking the area under the curve using the DescTools package in R^48^. Because of the considerable uncertainty around the R, we conduct a sensitivity analysis, calculating outbreak size over the whole range of estimated R values and the range of serial interval estimates used in the agent-based model.

### Agent-Based Simulation of Outbreak Size

#### Model Overview

We developed a discrete-time, metapopulation agent-based model (ABM) of measles transmission to estimate outbreak size in the affected community^42^. We implemented the model using the measles R package (version 0.3.2-0) with the epiworld C++ backend and conducted analyses in R (version 4.5).

#### Disease Model Structure

We use a modified Susceptible-Exposed-Infected-Recovered (SEIR) model (Figure SI 3) in which susceptible individuals (S) become exposed (E) following effective contact with an infectious individual. Exposed individuals progress to a prodromal infectious stage (Iₚ), representing the infectious period prior to rash onset, followed by an infectious rash stage (I ). Infected individuals either recover directly (R) or progress to hospitalization (H) prior to recovery. Transition rates between states were parameterized based on the natural history of measles. Table SI 1 shows the simulation parameters:

**Figure SI3:**
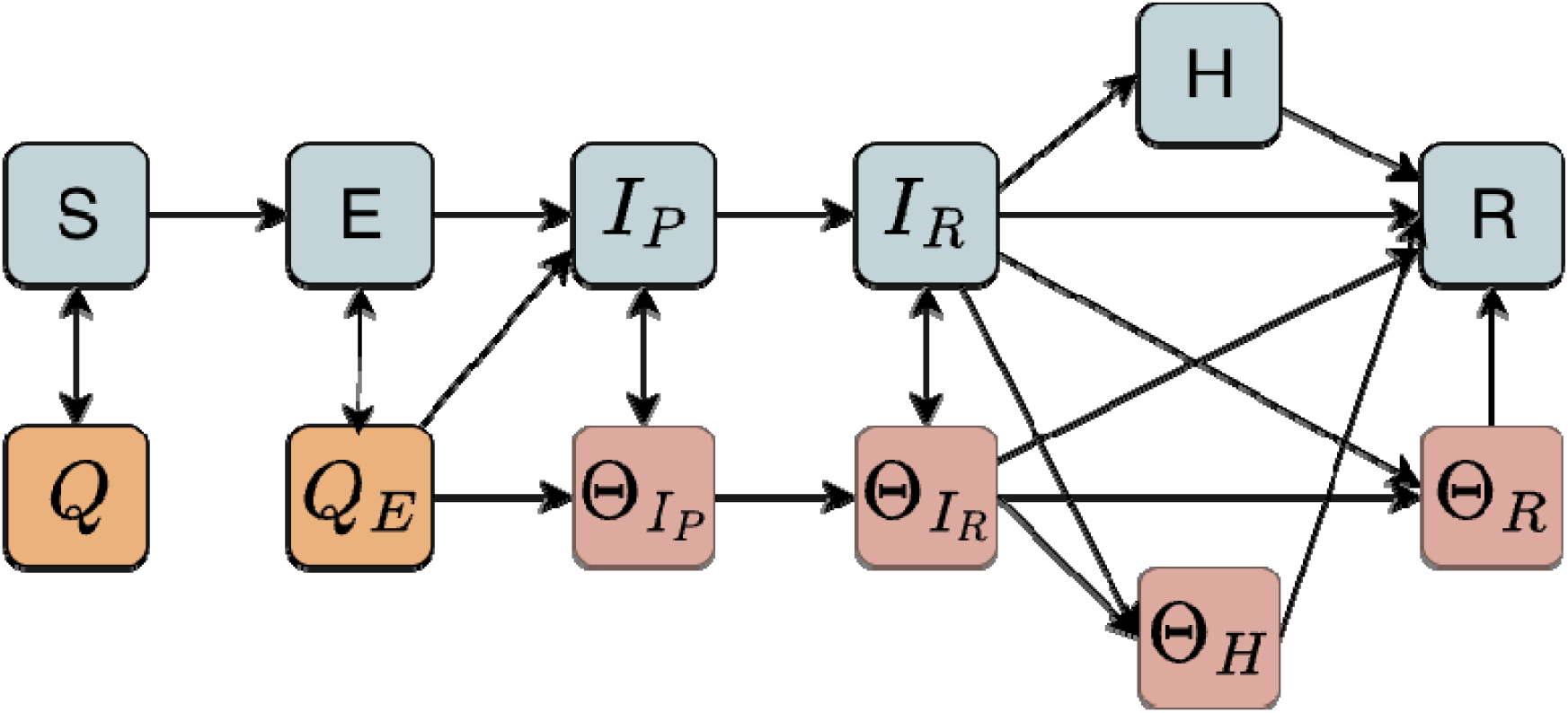
Model schematic of the measles ABM. The blue shaded boxes indicate the disease progression model with susceptibles (S) becoming exposed (E) progressing to infectious and prodromal (I_P_), then becoming infectious with a rash (I_R_) finally infections are either hospitalized (H) before they recover (R) or recover directly without hospitalization. The orange shaded boxes indicate quarantine (Q and Q_E_) and the red shaded boxes indicate isolated infections (□_IP_, □_IR_, □_H_, and □_R_). Quarantined susceptibles (Q) can only leave quarantine whereas quarantined exposed infections (Q_E_) can either leave quarantine or progress through the disease states while in isolation. Isolated infections similarly can leave isolation or progress through the disease states. Agents move between states at different rates depending on their vaccination status^49^. The model is stochastic except for the number of days agents spend in quarantine (21 days) and isolation (4 days).

#### Model Parameterization

##### Population Data

Population estimates for the Utah-Arizona border outbreak were obtained from the 2024 American Community Survey (ACS) 5-year estimates published by the U.S. Census Bureau^50^. We modeled a population of 5,913 individuals, representing the estimated size of the community.

##### Vaccination Data

School-level vaccination data were obtained from the Utah Department of Health and Human Services and the Arizona Department of Health Services. For the school in Utah, we tallied 2-dose MMR coverage data for elementary school, middle school, and high school levels as reported to the Utah Department of Health and Human Services, available via a public Shiny application^51^; and for Arizona schools, we tallied publicly available online data^14^.

##### Age / school group population size and immunization assumptions

Based on census data and school size estimates for different ages corresponding to school levels, we organized the community into 15 groups, consisting of 9 distinct schools and 6 groups comprising non-school-age community members of different age groups. For the age-0 group, we assumed no vaccination coverage as the first MMR dose is not recommended until 12 months of age. For the age 1-4^39^ group, we assumed the same 1-dose MMR coverage as the average of the 2-dose coverages of the three elementary schools. For the 9 school groups, we estimated the vaccination coverage directly from the data described above. For the age 18-24 group, we assumed coverage equal to the average of the high schools. For the age 45-69 group we assumed 95% coverage, and for the age 24-44 group we assumed the average of the 18-24 and the 45-69 groups. We assume 97% vaccine effectiveness of two-dose MMR for ages 5+, and 93% vaccine effectiveness of one-dose MMR for ages 1-4. For those aged 70 and older, we assumed 100% immunity due to childhood infection with measles before vaccination was available.

**Table SI 1:** Simulation Parameters for the Agent Based Model

| Parameter | Meaning | Value |
| --- | --- | --- |
| Contact tracing window | How many days before/after rash onset will be considered for the contact tracing | 4.00 days |
| Contact tracing success rate | The average proportion of cases where a contact will be detected, per event | 0.25 |
| Mean days undetected | Average number of days from rash onset to detection | 2.00 days |
| Mean Hospitalization period | Average number of days an infection is hospitalized | 7.00 days |
| Hospitalization rate | Adjusted to match a 10% hospitalization probability | 0.037/day |
| Mean incubation period | Average duration of time from exposure to prodromal symptom onset | 12.00 days |
| Isolation period | Fixed duration of time that infections with rash are isolated | 4.00 days |
| Isolation willingness | Probability of adherence to isolation | 0.90 |
| Mean prodromal period | Average duration of non-specific symptoms | 4.00 days |
| Quarantine period | Fixed duration of time that contacts of infected individuals are quarantined | 21.00 days |
| Quarantine willingness | Probability of adherence to quarantine | 0.90 |
| Mean rash period | Average duration of rash symptoms | 4.00 days |
| Vaccine efficacy | Effectiveness of the vaccine at mounting an immune response, that is, 97% of vaccinated individuals are totally immune and 3% are totally susceptible | 0.97 |

##### Population Structure and Mixing

We use a metapopulation or structured population framework for our ABM, and partition agents into subpopulations based on school and age groups. We assigned school-aged individuals to specific schools and grouped non-school-aged individuals by age based on US Census demographic distributions. These subpopulations defined the structure through which transmission occurred.

We generated an age- and school-structured contact matrix using our contactMatrixAgeSchool() function from the multigroup.vaccine R package^19^. The function first creates a baseline age-specific contact matrix from social contact survey data via the socialmixr R package^52^, which derives symmetric age-stratified contact matrices from empirical diary-based contact data from the POLYMOD study^31^. We adjusted the symmetric POLYMOD-derived contact matrix to the Arizona-Utah border population by dividing out the combined population age distribution of all the POLYMOD countries at the time the surveys were administered and replacing it with the age distribution of our modeled population. We then subdivided the school-aged groups into separate school-specific subgroups while preserving the overall age-based mixing structure.

The contactMatrixAgeSchool() function redistributes between-age-group contacts involving a school-age group across its school subgroups in proportion to school population sizes. The function models within-school-age-group contacts using a preferential and proportional mixing function, applying a preferential within-school mixing value of 70%, representing the fraction of within-age-group contacts occurring exclusively within the same school. Specifically, following established methods^53^, 70% of each school group’s within-age-group contacts are assigned exclusively to its own school, and the remaining 30% are distributed among all schools, including its own, in proportion to the school sizes, representing random within-age-group contacts outside of school hours. The resulting matrix preserved overall age-specific contact rates while explicitly modeling enhanced assortative mixing among individuals attending the same school and while ensuring contact symmetry over the population.

##### Transmission and Calibration

We modeled transmission as a frequency-dependent process and calibrated the overall contact rate to reproduce R_0_=9. We assigned vaccination coverage using school-level data for school-aged individuals as detailed above.

##### Public Health Interventions

We explicitly modeled isolation of confirmed cases and quarantine of identified contacts as independent public health interventions, each triggered following case detection, which was assumed to occur after rash onset with an average delay of two days (geometric distribution). Isolation and quarantine were parameterized separately, with adherence to each varying independently across scenarios. We explored all combinations of 0%, 25%, 50%, 75%, and 100% adherence for both isolation and quarantine, yielding 25 scenarios in total; results for three scenarios are presented in the main text and all remaining combinations are reported in the supplement. Quarantined susceptibles may only leave quarantine, whereas quarantined exposed individuals may either leave quarantine or progress through disease states while in isolation; isolated individuals may similarly leave isolation or continue progressing through disease states. Quarantine and isolation durations were fixed at 21 and 4 days, respectively; all other transitions were modeled stochastically

##### Simulation Design

We initialized simulations with a single infectious individual and ran 1000 stochastic simulations, each for 365 days, to capture the full outbreak trajectory. We defined total outbreak size as the cumulative number of infections over the course of each simulation.

##### R_0_ Sensitivity Analysis

Given the variability in estimates of the basic reproduction number of measles and the substantial impact of R_0_ on outbreak dynamics, we conducted a sensitivity analysis across a range of R_0_ values calculated from our estimates of Rₜ to assess the robustness of our results to assumptions about transmissibility. Although the basic reproduction number for measles is typically reported to range from 12 to 18^3^, we evaluated a broader range (R_0_ = 5-17) to reflect characteristics of the Utah outbreak and to encompass biologically plausible lower transmission scenarios. This range was informed by estimates of the effective reproduction number in Utah during the early outbreak period.

For each value of R_0_, we recalibrated the transmission parameter in the agent-based model to achieve the target reproduction number under baseline conditions. All other model parameters, including population structure, vaccination coverage, contact patterns, and intervention assumptions, were held constant. We repeated the simulation procedures described in the primary analysis for each R_0_ value.

##### Population Structure and Mixing Sensitivity Analysis

Although the POLYMOD study is a foundational study on human contact patterns including age-assortative mixing and setting-specific interactions (e.g., schools), the POLYMOD study is geographically limited and was conducted prior to the COVID-19 pandemic when mixing patterns changed^54^. To assess the robustness of our findings to assumptions about social mixing, we conducted a sensitivity analysis in which the contact structure of the agent-based model was replaced with empirical age-stratified contact matrices from the Epistorm-Mix study^55^, a probability-based survey of 1,930 respondents designed to be nationally representative of the post-pandemic U.S. population by age, sex, race/ethnicity, household income, census region, and language. To enable a fair comparison, the per-contact transmission probability was recalibrated for each mixing scenario so that the basic reproduction number R_0_ matched the target value used in the main analysis (R_0_ = 9) and for each of the R_0_ values used in the R_0_ sensitivity analysis. This ensures that any differences observed between scenarios reflect the structure of mixing rather than its overall intensity.

The contact matrix was first adjusted to reflect the age distribution of the study region. Because our model treats individual schools as distinct population blocks, we further modified contact rates for school-age individuals, assuming that 70% of contacts among school-age agents occur within their own school. Overall contact rates were preserved but redistributed to reflect this within-school concentration.

Since the Epistorm contact matrix uses five-year age intervals (0-4, 5-9, 10-14, etc.), we mapped it to our model’s age groups using a weighted average. For any model age group spanning multiple Epistorm intervals, the effective contact rate was computed as a weighted average across overlapping intervals, with weights proportional to the number of ages from each interval represented in the model group. We ensure to apply the same transformations as with the POLYMOD data, particularly adjusting it to the age distribution and applying the preferential within-school mixing value of 70%, while preserving symmetry.

## Author contributions

A.M., D.J.A.T., J.W., and L.T.K. contributed to data curation. A.M., D.J.A.T., G.V.Y., J.W., and L.T.K. performed formal analysis. D.J.A.T., E.T.L., M.H.S., L.D.N., A.T.P., K.O., and L.T.K. contributed to conceptualization and study design. A.M. and L.T.K. created visualizations. A.M., J.W., and L.T.K. wrote the original draft. All authors reviewed and edited the manuscript. L.D.N., K.O., and L.T.K. provided supervision and project administration. E.T.L., M.H.S., and L.T.K. acquired funding.

## Competing Interests statement

The authors declare no competing interests.

## Funding Statement

This publication was made possible by cooperative agreement CDC-RFA-FT-23-0069 from the CDC’s Center for Forecasting and Outbreak Analytics. Its contents are solely the responsibility of the authors and do not necessarily represent the official views of the Centers for Disease Control and Prevention.

DJAT, GVY, JW, ETL, MHS, ATP, LDN, and LTK disclose support for the publication of this work from the CDC’s Center for Forecasting and Outbreak Analytics [cooperative agreement CDC-RFA-FT-23-0069].

AM and KO disclose support for the research of this work from the Epidemiology and Laboratory Capacity (ELC) program at the CDC Cooperative Agreement Number CK24-0002

ETL discloses support for the research of this work from the National Institutes of Health (R35GM147013).

## Data availability statement

Measles whole-genome sequences are deposited in the NCBI Sequence Read Archive under BioProject accession PRJNA1293457. Reported case data used for R_t_ estimation are publicly available from the Utah measles dashboard. Vaccination coverage data were obtained from the U.S. Census Bureau American Community Survey and from publicly available school-level reports from the Utah Department of Health and Human Services and the Arizona Department of Health Services. Individual-level case and contact tracing data cannot be shared due to patient confidentiality

## Code availability statement

All code used for phylodynamic analysis and agent-based modeling is publicly available at https://github.com/EpiForeSITE/measles-hidden-burden

## Notes

### Competing Interest Statement

The authors have declared no competing interest.

